# MRI-based surface reconstruction and cortical thickness estimation of the human brain: Benchmarking deep-learning based morphometry tools

**DOI:** 10.1101/2025.03.22.25324274

**Authors:** Victor B. B. Mello, Richard McKinley, Roland Wiest, Christian Rummel

## Abstract

Establishing reliable and time efficient pipelines for structural MRI segmentation, parcellation and surface reconstruction, is essential to explore the potential clinical applications of research-grade morphometry tools. The integration between deep-learning based methods for fast whole-brain segmentation and the well known surface reconstruction algorithms is a viable alternative to perform this task. In this work, we applied this idea with three deep-learning based cortical parcellation models, DeepSCAN, FastsurferCNN and QuickNAT. With a 11 min surface reconstruction pipeline, we evaluated the performance of each segmentation beyond the voxel-based approaches and dice coefficient comparison between the generated parcellation and Freesurfer’s established silver standard. To prove the concept, we performed a direct comparison between the morphological variables obtained by our methodology and Freesurfer. Using a synthetic dataset, we benchmark each reconstruction pipeline based on the similarity to the ground-truth surface and reproduction of the expected surface-based metrics. The most robust pipeline across the human dataset and closer to the synthetic ground truth was based on DeepSCAN segmentation, producing a reliable morphometric tool with a processing time realistic for clinical applications like diagnostics support in individuals.

## Introduction

Quantitative analysis of structural Magnetic Resonance Imaging (MRI) has been available for more than 20 years [1–3], serving as a powerful tool for studying the healthy brain and different neurological disorders across the human lifespan [4]. Whole-brain reconstruction from a structural MRI is typically performed by time consuming processing by one of a handful of standardly accepted tools: voxel-based morphometry (VBM), as provided by SPM [5], ANTs [6], and FSL [7], or cortical surface reconstruction for surface-based morphometry (SBM), as performed by Freesurfer [8], BrainSuite [9] and CIVET [10]. The complexity of the cortical geometry, the high biological variability across subjects and the memory requirements of the data are the main reasons that whole-brain reconstruction is so computationally demanding.

From a clinical perspective, structural MRI provides a non-invasive visual assessment of the brain, with well established protocols for diagnosing and monitoring a wide range of neurological conditions [11–13]. Furthermore, various forms of brain morphometry derived from these images have significantly contributed to a deeper understanding of healthy brain development, aging, and the mechanisms underlying disease manifestation. For this reason, it would be beneficial to translate the morphometry tools produced within the research context to auxiliary quantitative assessments that can used in clinical practice [14–17]. In order to achieve this goal, it is essential to devise new reconstruction pipelines that are not only capable of fully reconstructing a structural MRI in a time efficient manner but also are robust and accurate against the inherent variability in the reality of clinical applications, e.g., multiple MRI machines, different imaging contrast and subjects.

Deep-learning (DL) [18] based methods have become the first choice for fast whole-brain segmentation and parcellation. In the last few years, the field has rapidly evolved from architectures that generate brain segmentation with a runtime of 15-60 min [19–24] to models with a runtime of less than a minute [25–28]. However, there is much progress to be made in the use of DL for brain surface reconstruction. Only a few models are available [29–31] and, although they can be very fast (less than a 1 min runtime), they still generate surfaces with self-intersecting regions. The state-of-the-art model available [31] reported an improvement on the accuracy and regularity of DL-based surfaces, with a reduction from (1.47 *±* 0.52)% to (0.24 *±* 0.17)% of self-intersecting faces on the reconstructed pial. Still, there is no guarantee that the reconstructed surfaces will have the required topological properties. This makes Fastsurfer [28], which reconstructs a cortical surface by combining DL-based segmentation and Freesurfer in about 90 min, the fastest whole-brain reconstruction pipeline described in the literature so far with the certainty of generating topologically correct surfaces.

The advent of novel structural MRI reconstruction techniques makes the development of methods to probe their accuracy imperative. However, a major obstacle to such evaluation is the lack of datasets with their indisputable ground truth reconstruction. The manual production of such golden standard by experts [32] is a time-consuming task that requires highly-specialized anatomical knowledge and may present significant variability across different experts [33]. To overcome this problem using real structural MRI, it is common to use Freesurfer reconstructions as a silver standard given the long standing trustworthiness on its results. An alternative is using multiple MRI acquisitions of the same cortex to perform a test-retest reliability assessment [28, 34]. There are some caveats, with the former leading to an evaluation bias [34] and the latter being only a indirect indication of reconstruction accuracy. Another possible solution is the use of a synthetic ground-truth dataset generated by a controlled modification on a real brain MRI. As cortical thickness is a metric routinely used to study neurodegenerative and neurological conditions, this approach typically focuses on inducing synthetic atrophy by shrinking the cortex. By producing a topology-preserving deformation field from the pial surface to the white matter (WM) – grey matter (GM) interface, the atrophy is generated by moving the pial along this field, in the direction of the WM. The available models to create a synthetic cortical atrophy can be based on the Jacobian of the transformation [35–37], biomechanical models [38–40], binary morphological operations [41] or DL [42, 43].

To extend the analysis of the relative performance of DL-based segmentation tools beyond the dice coefficient between the alternative cortical parcellation and a silver standard ground truth as provided by Freesurfer, in this work we aim to examine also the quality of morphometric variables extracted from these segmentations. To maximize compatibility with fast clinical deployment, we developed a fast surface reconstruction pipeline deeply inspired by the approach presented in [28]. This yields a reliable pipeline that can be applied to any high-quality segmentation method, reducing processing time for whole-brain reconstruction down to minutes. In this work we evaluate three state-of-the-art DL-based tools for whole-brain MRI segmentation for morphometry, DeepSCAN [27], FastsurferCNN [28] and QuickNAT [26] To prove the concept, we performed a direct comparison between the morphological variables obtained from DL-based segmentations by our proposed fast surface reconstruction pipeline and Freesurfer for a wide variety of subjects and MRI sequences. Then, using the synthetic dataset generated by [43], we benchmarked each reconstruction pipeline based on the similarity to the ground-truth surface and reproduction of the expected surface-based metrics.

## Materials and Methods

### Imaging data

We study morphometric pipelines applied to both real T1w human brain MRI (obtained from open-access databases) and synthetic data (the DL-based cortical atrophy phantom generated by [43]). The human dataset aims to provide a proof of concept for the fast surface reconstruction pipeline using DL-based whole-brain segmentation. In order to demonstrate the generalizability of the fast surface methodology, the human dataset was compiled with a high variability regarding age, pathology and MRI scanning parameters for both male and female subjects. The dataset combines: (i) a random sample of 1000 young subjects from the Adolescent Brain Cognitive Development (ABCD) study [44]; The healthy young adults from the Human Connectome Project (HCP) [45]; (iii) The Phantom of Bern (PoB) [46], a longitudinal experiment with two healthy adults in which the MRI sequence parameters inversion time (TI) and repetition time (TR) are systematically changed to control the contrast between the WM and GM signal intensities; (iv) The elderly subjects, both healthy and with some degree of cognitive impairment, from the OASIS3 dataset [47]. A summary description of the human MRI dataset is given in Table 1. Both HCP and OASIS3 dataset have subjects with more than one MRI scan taken at the same session, allowing a reproducibility test-retest analysis. With the exception of the PoB, which was designed for a controlled change on the MRI scanning parameters, all datasets were obtained in studies with more than one scanner and distinct acquisition parameters.

**Table 1:** Summary of the human MRI dataset. Abbreviations: CTL, healthy control, AD, Alzheimer’s disease.

| Datasets | Diagnosis | N subjects |  | N scans |  | Age range [years] | Scanners |
| --- | --- | --- | --- | --- | --- | --- | --- |
|  |  | Male | Female | Male | Female |  |  |
| ABCD | CTL | 500 | 500 | 500 | 500 | 9-10 | 3 |
| HCP | CTL | 507 | 606 | 815 | 968 | 26-35 | 3 |
| OASIS3 | CTL | 299 | 438 | 777 | 1226 | 42-97 | 3 |
| OASIS3 | AD | 193 | 167 | 338 | 298 | 50-95 | 3 |
| PoB | CTL | 2 | 0 | 18 | 0 | 30’s and 40’s | 1 |

The synthetic dataset was generated using 20 heathy controls from the Alzheimer Disease Neuroimaging Initiative (ADNI) [48, 49] by exploiting the inherent one-to-one correspondence between vertices of the WM surface and vertices of the pial surface on Freesurfer’s reconstructed meshes. A uniform controlled atrophy across the GM was introduced moving the pial surface in the direction of the WM with a fixed distance. Then, based on these deformed meshes, a synthetic MRI was generated using a generative adversarial network (GAN) model [50]. The synthetic dataset provides an expected value for the cortical thickness and ground-truth surfaces for the pial and white matter, allowing a direct comparison to benchmark each reconstruction methodology. The values of atrophies provided by the authors range from zero (no modification of the surfaces) to 1 mm.

### The processing pipeline with different segmentation/surface reconstruction methods

All datasets were first processed using (A) Freesurfer v6.0 [8] reconstruction pipeline following the Desikan Killiany (DK) protocol atlas parcellation [51]. No manual corrections were performed [52]. These reconstructions for the human dataset were used as a silver standard comparison baseline given the long standing reliability of Freesurfer reconstructions (despite the inherent bias towards Freesurfer-based methods)

For reconstruction from DL-based segmentation, we focused on three different methods: (B) DeepSCAN [27], (C) FastsurferCNN [28] and (D) QuickNAT [26]. FastsurferCNN and QuickNAT can be regarded as variants of the same architecture, with the former being inspired by the later. All human datasets were processed by the three DL-based segmentation models. Since (D) QuickNAT cannot process skull-stripped imaging, it was not applied to the synthetic CTh phantom.

The surfaces for each DL-based segmentation method can be obtained with minimal modifications on the pipeline presented in [28]. Given a proper conformation of the input segmentation and a labeling map consistent with the number of labels that each DL model can predict, the surfaces are obtained with a significant speed-up compared to Freesurfer. Heavily inspired by this, the fast surface pipeline suggested in this work is a simplified version of the one suggested by [28]. Table 2 summarizes the steps omitted to produce the fast surface reconstruction pipeline suggested here. Based on the segmentation provided by each DL model, the WM surface is reconstructed using a topology correcting fast marching algorithm [53]. This surface encloses the cerebral WM and sub-cortical structures such as the ventricles, thalamus, caudate nucleus, putamen, pallidum, hippocampus, amygdala, accumbens nucleus and ventral diencephalon. Only the pial surface is reconstructed using the Freesurfer engines, requiring functionalities that normalize the MRI to ensure a WM signal with about 110 in gray-scale, modify the WM segmentation to avoid edges or corners and position the tessellation of the cortical surface [1]. Finally, each vertex of the tesselation is mapped on the labels provided by the segmentation with no spatial smoothing applied on the labels, allowing a region-wise analysis.

**Table 2:** Summary of the modifications made to the Fastsurfer reconstruction pipeline.

| Fastsurfer Processing [28] | Fast pipeline (This work) |
| --- | --- |
| 1. DL-based parcellation | yes, according to methods (B), (C) or (D) |
| 2. Bias field correction | no |
| 3. Linear Talairach registration | no |
| 4. Intensity normalization | yes |
| 5. Corpus-Callosum segmentation | no |
| 6. Remove edges or corners in WM segmentation | yes |
| 7. WM tessellation using Marching Cubes | replaced by topology correcting fast marching [53] |
| 8. Spherical mapping | no |
| 9. Fix topology | no |
| 10. Place WM surface | no |
| 11. Make pial surface (deform WM surface) | yes |
| 12. Map to cortical labels | yes |

To ensure that our simplification of the Fastsurfer pipeline does not affect the surfaces and results presented in this paper, the full Fastsurfer pipeline was used to clarify cases with large discrepancy in comparison to Freesurfer. These results are summarized in the supplementary material A. All the codes necessary to run the fast pipeline reconstruction are available at https://github.com/bragamello. The performance achieved in this paper was obtained using a computer with 8 Intel Xeon(R) E5 1630 v3 3.70 GHz Central Processing Unit (CPU) and a 8 Gb GeForce GTX 1080 Graphical Processing Unit (GPU).

### Data analysis

From each reconstruction, we derived five common surface-based metrics: the volume enclosed by the WM and pial surfaces, their respective surface areas, and the cortical thickness. Cortical thickness was calculated from the reconstructed WM and pial surfaces using Freesurfer’s definition [54]. This method averages the closest distance measured from WM to pial surface and from pial to WM surface to generate a vertex-wise thickness estimate. The morphological variables were compared both by their values per hemisphere and regionally according to the DK atlas. In the case of FastsurferCNN predictions, the cortex parcellation follows the Desikan-Killiany-Tourville (DKT) Atlas [55]. To account for this difference, the bankssts, temporal pole and frontal pole were omitted in our region-wise comparisons. Given the small size of these regions, the effect of this difference has no impact on our general observations. As the synthetic dataset was conceived to benchmark thickness estimation methods, we included comparisons with a registration-based cortical thickness measurement (DiReCT) [56] for this particular dataset. This methodology not only was suggested [27] to detect subtle changes in cortical thickness from a MRI together with their segmentation model but also showed good performance with the synthetic dataset [43]. Table 3 summarizes all the processing pipelines and datasets available in this work.

**Table 3:** Summary of the processing pipelines used in each dataset. With the exception of QuickNAT, all models produce a brain parcellation allowing the assignment of vertex-wise labels to the pial surface reconstruction.

|  | (A) Freesurfer |  | (B) DeepSCAN |  | (C) FastsurferCNN |  | (D) QuickNAT |  |
| --- | --- | --- | --- | --- | --- | --- | --- | --- |
|  | Parcellation | Thickness | Parcellation | Thickness | Parcellation | Thickness | Parcellation | Thickness |
| ABCD | yes | FS | yes | FS | yes | FS | no | FS |
| HCP | yes | FS | yes | FS | yes | FS | no | FS |
| OASIS3 | yes | FS | yes | FS | yes | FS | no | FS |
| PoB | yes | FS | yes | FS | yes | FS | no | FS |
| Synthetic | no | FS/DiReCT | no | FS/DiReCT | no | FS/DiReCT | – | – |

FS = Freesurfer’s thickness; DiReCT = Diffeomorphic registration based cortical thickness measurement

For whole-hemisphere comparisons, we performed linear regression on the surface-based metrics derived from Freesurfer and our alternative method to estimate the slope and offset with a 95% confidence interval (C.I.). In the region-wise analysis, we calculated the median percentage deviation of the regional surface-based metrics relative to Freesurfer values. Additionally, to evaluate test-retest reliability, we used the repeated MRI scans from the HCP dataset. Using all possible combinations of repeated scans, we calculate de root mean square (RMS) of the difference in the surface-based metrics, reporting the mean value for each region. Although repeated MRI scans were also available for OASIS3, the cohort with young healthy adults were preferred in this analysis to avoid introducing extra confounder, such as repeated subjects due to bad quality images or extreme cases of atrophy. The relationship between surface reconstruction and the contrast between the WM and GM signal intensities on the MRI was examined using the PoB dataset by determining the correlation between all surface-based metrics and the measured WM/GM contrast. Following Freesurfer’s standard, the local WM/GM contrast was calculated based on the reconstructed WM surface, according to the ratio 2 *×* (*I*_WM_ *− I*_GM_)*/*(*I*_WM_ + *I*_GM_), by sampling *I*_WM_ tissue intensity values at 1 mm inside the WM and *I*_GM_ reaching 30% of the cortical thickness inside the GM. Regarding the synthetic dataset, the similarity of the surface reconstructions to the ground-truth surface was computed using the Hausdorff distance and the relative error in the measurement of the surface areas and volumes. For benchmarking the thickness estimation methodologies, we directly compared the synthetically imposed atrophy with the estimated one.

## Results

### Surface reconstruction

A general overview of the surfaces obtained by the fast pipeline and the cortical parcellation provided by its respective DL-based model is given in Figure 1 using a scan from the PoB dataset with a mid-value contrast as an example. Further examples can be found at the Supplementary Material 12

**Figure 1:**
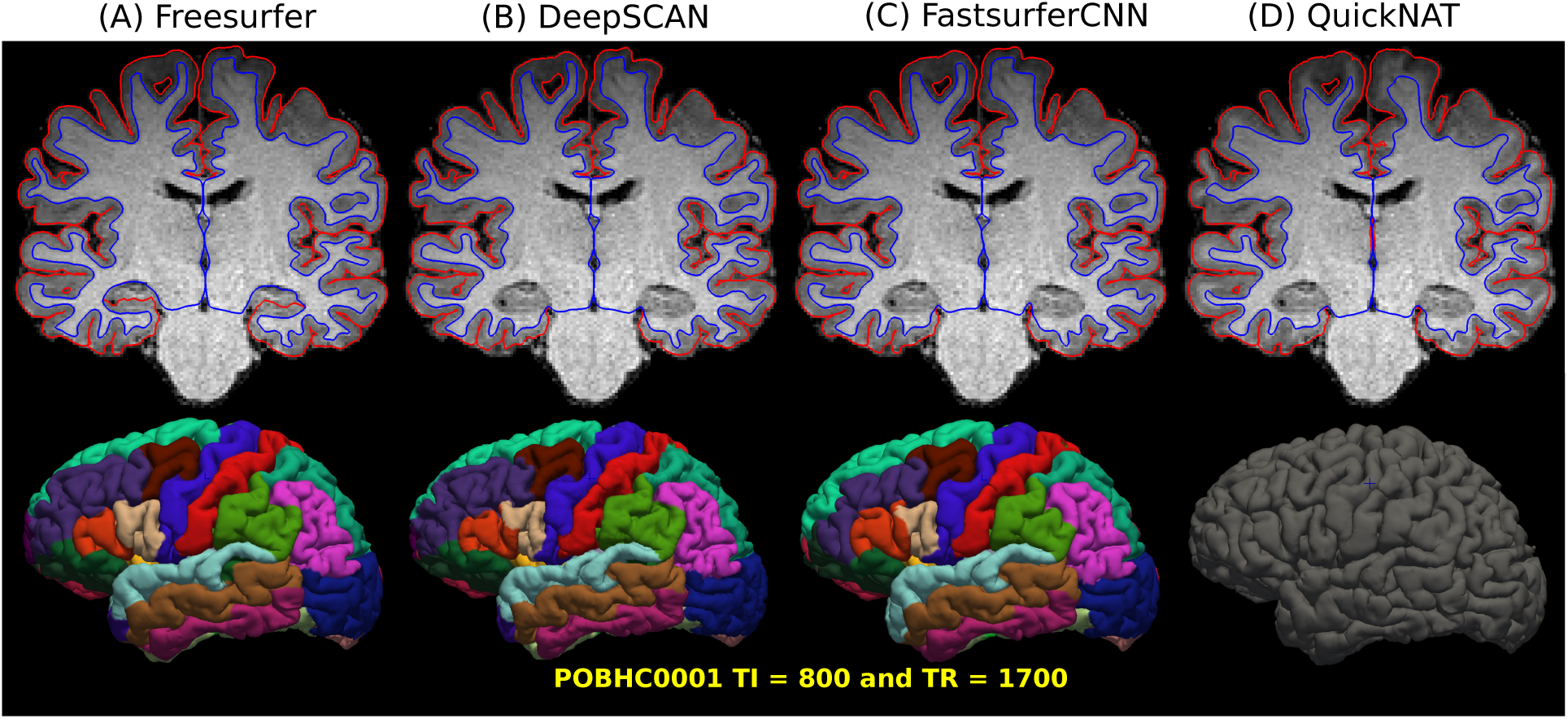
Example of the results obtained from the fast surface reconstruction pipeline based on (B) DeepSCAN, (C) FastsurferCNN and (D) QuickNAT segmentation in comparison with the standard (A) Freesurfer reconstruction. The raw MRI is displayed together with the reconstructed WM (blue) and pial surfaces (red) for the same coronal slice to facilitate a direct comparison between surfaces. Apart from occasional segmentation failures, the surfaces are similar. A systematic exception is the hippocampus, which is not enclosed inside the WM for (A) Freesurfer. The 3D reconstruction of the pial surface showing the region labeling provided by each DL-based segmentation model allows a neuro-anatomical comparison with the DK Atlas. The subject from the PoB dataset is identified for reproducibility and the anatomical visualization was kept consistent to facilitate comparison between reconstructions. We retained the same coloring and naming presented in [51].

The distribution of runtimes required to reconstruct a healthy young adult structural MRI with the fast surface pipeline is shown in Figure 2 with a direct comparison with Fastsurfer and Freesurfer. Importantly, our fast surface pipeline is a simplification of Fastsurfer, which generates only the surfaces, the vertex-wise thickness, curvature and annotation as output but not the many other metrics that FreeSurfer and FastSurfer provide. This runtime reflects the minimum amount of time required to generate a surface and minimal outputs that can be further manipulated according to the desired application.

**Figure 2:**
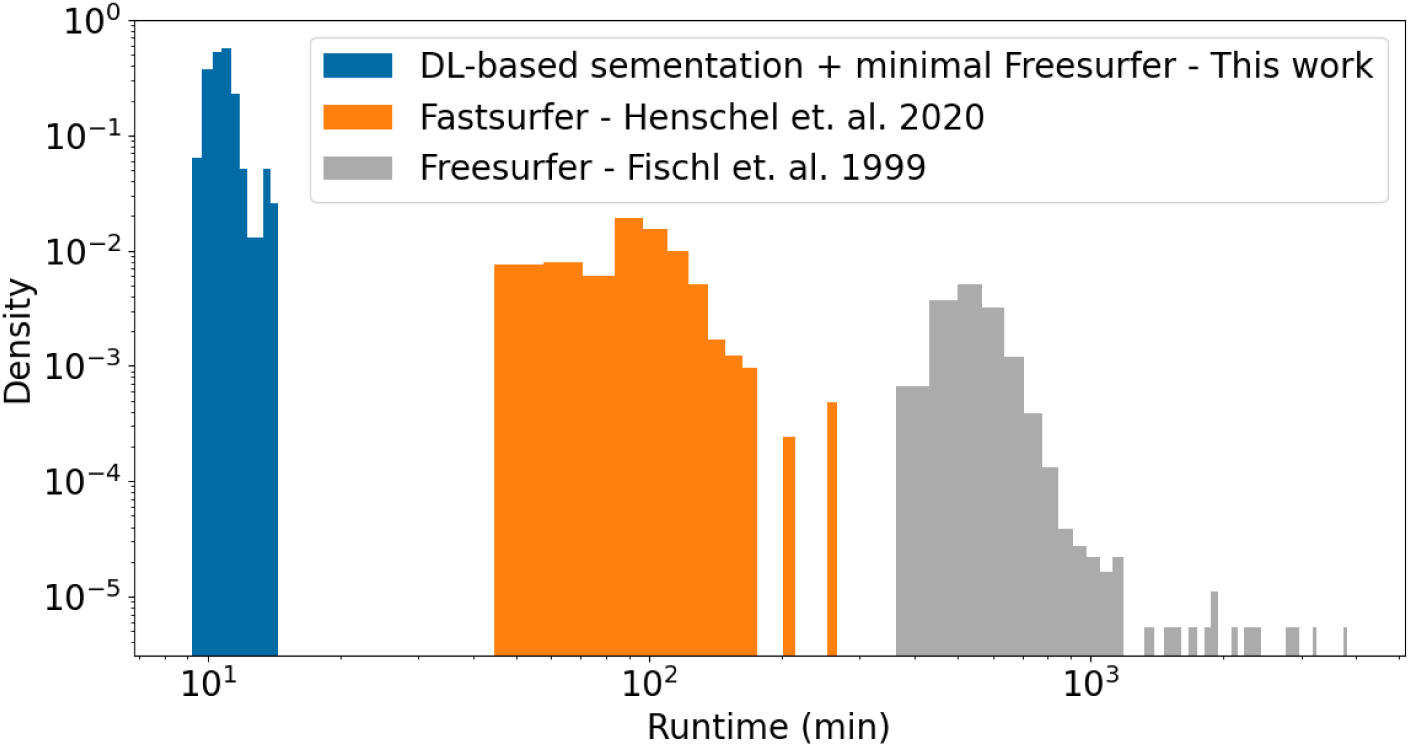
The runtime distribution for the fast surface reconstruction, Fastsurfer and Freesurfer pipelines. Due to the significant difference in the time scale from minutes to hours, the results are shown in log scale.

The analysis per hemisphere for subjects of the ABCD, HCP and OASIS3 datasets is shown in Figure 3. In the supplementary information C, the Table 5 summarizes the values of the slope and offset with 95% C.I. obtained by the linear regression between surface-based metrics based on Freesurfer and the alternative fast surface reconstruction method.

**Figure 3:**
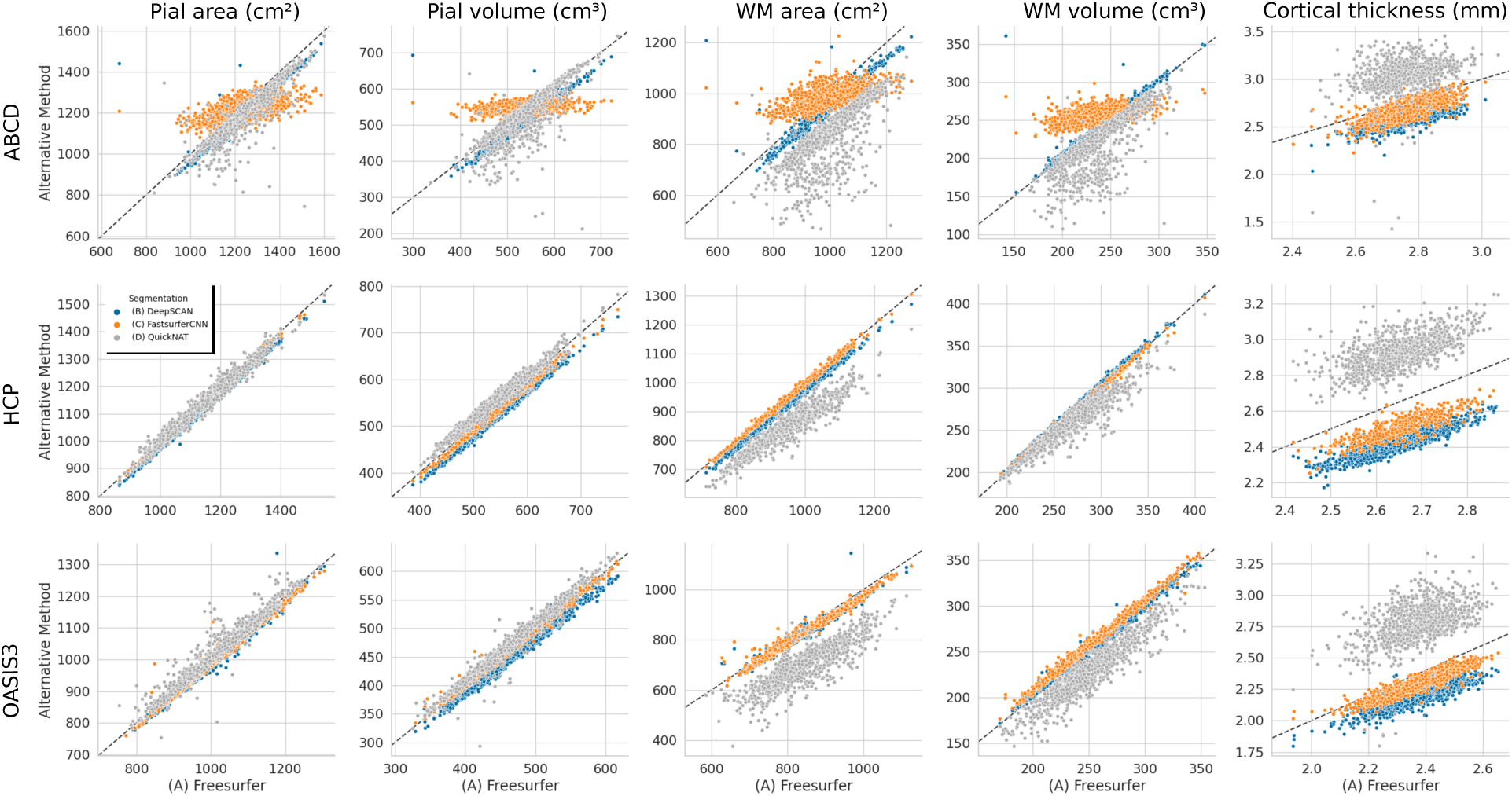
Comparison between the SBM metrics for a whole hemisphere obtained using the fast surface reconstruction based on (B) DeepSCAN, (C) FastsurferCNN and (D) QuickNAT segmentation models (y axes) and the standard (A) Freesurfer reconstruction (x axes). Each data point represents a subject, with the metric value obtained by the average between left and right hemispheres. The identity mapping is indicated by a dotted diagonal line.

The regional median relative difference to Freesurfer’s pial area and cortical thickness calculated region-wise is shown in Figure 4 following the DK atlas. Since (D) QuickNAT does not provide a parcellation of the cortex, method (D) is excluded here. A test-retest reliability assessment in HCP subjects with repeated scans is shown in Figure 5 by the mean RMS of difference in the metrics calculated using repeated MRI scans.

**Figure 4:**
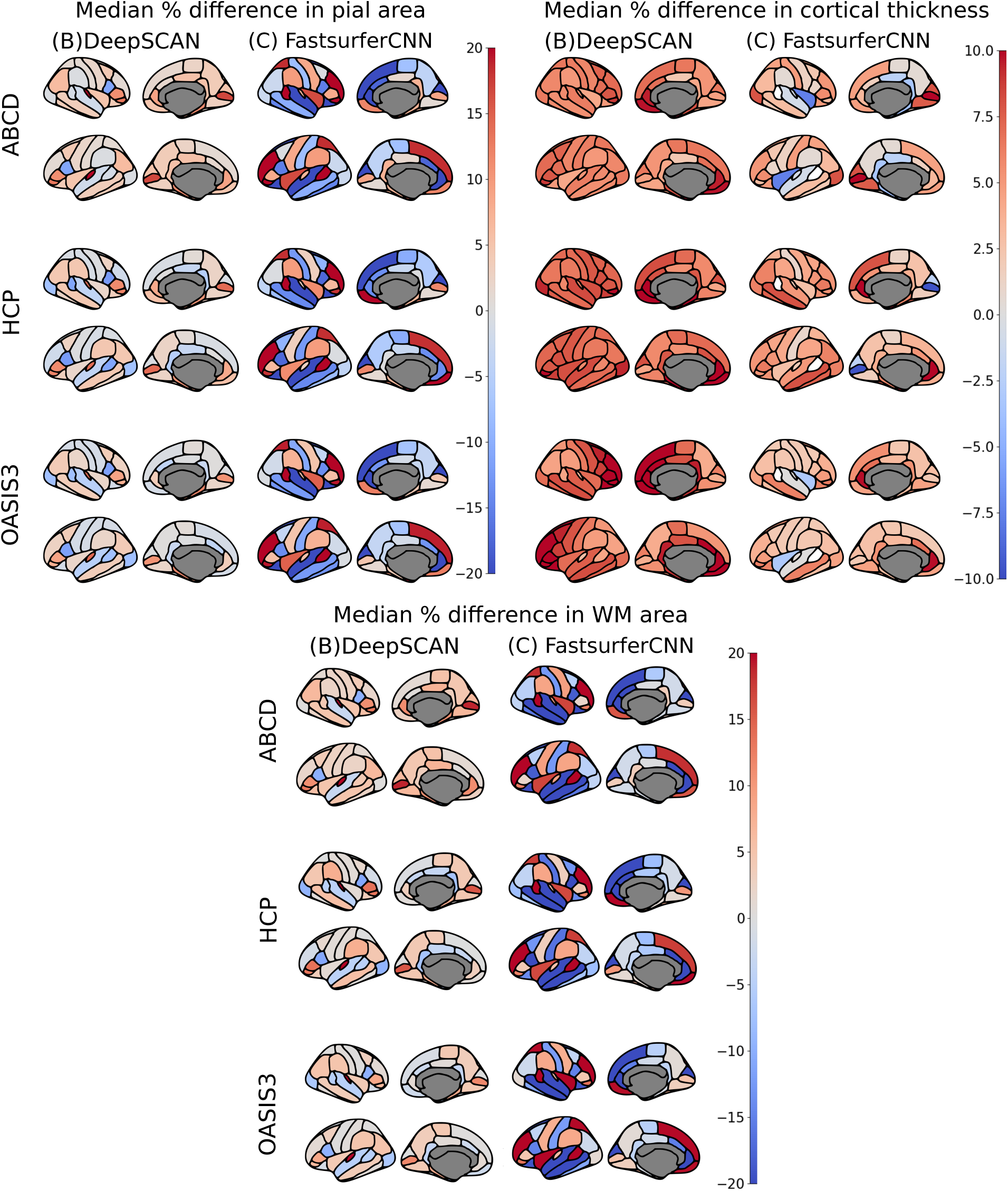
Median relative difference in the regional pial area, WM area and mean cortical thickness between Freesurfer reconstruction and the alternative fast surface reconstruction method based on (B) DeepSCAN, (C) FastsurferCNN segmentation. Negative values (blue hues) imply that values smaller than for Freesurfer were obtained. The DK atlas was used for parcellation.

**Figure 5:**
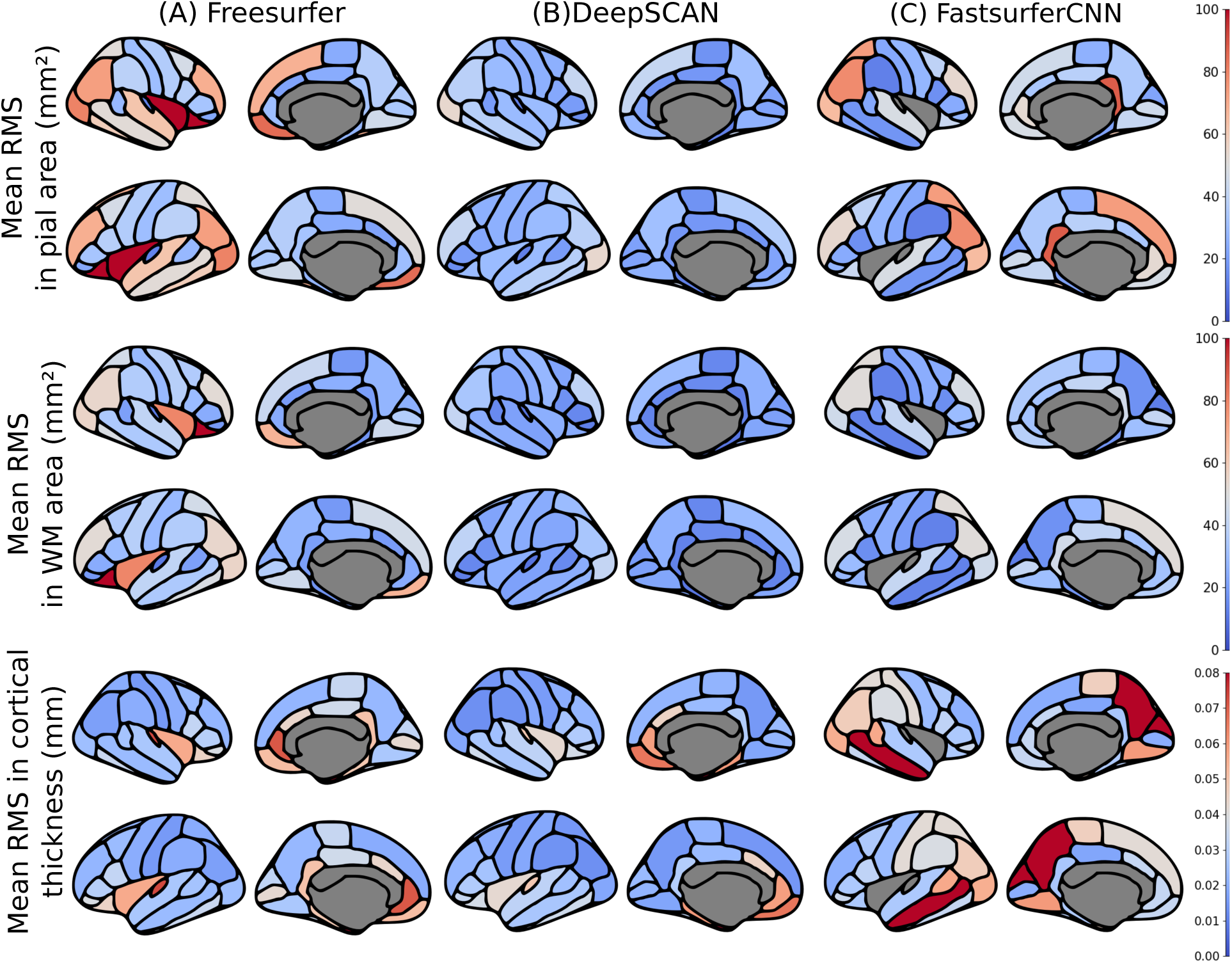
Test-retest reliability assessment of the pial surface area (top), WM surface area (middle) and cortical thickness (bottom) using HCP subjects with repeated MRI scans. Following the DK atlas, we determine the mean RMS of each metric using all rescan pairs of the same subject for (A) Freesurfer reconstruction and the fast surface pipeline based on DeepSCAN and (C) FastsurferCNN.

The impact of different MRI WM/GM contrast on the reconstructed surface and the derived surface-based metrics is shown in Figure 6 for the PoB dataset. The volumes enclosed by the pial and WM surfaces were used to determine if the surfaces are shrinking or increasing as a function of the contrast. Table 4 summarizes the correlation coefficient and the statistical significance of the results.

**Figure 6:**
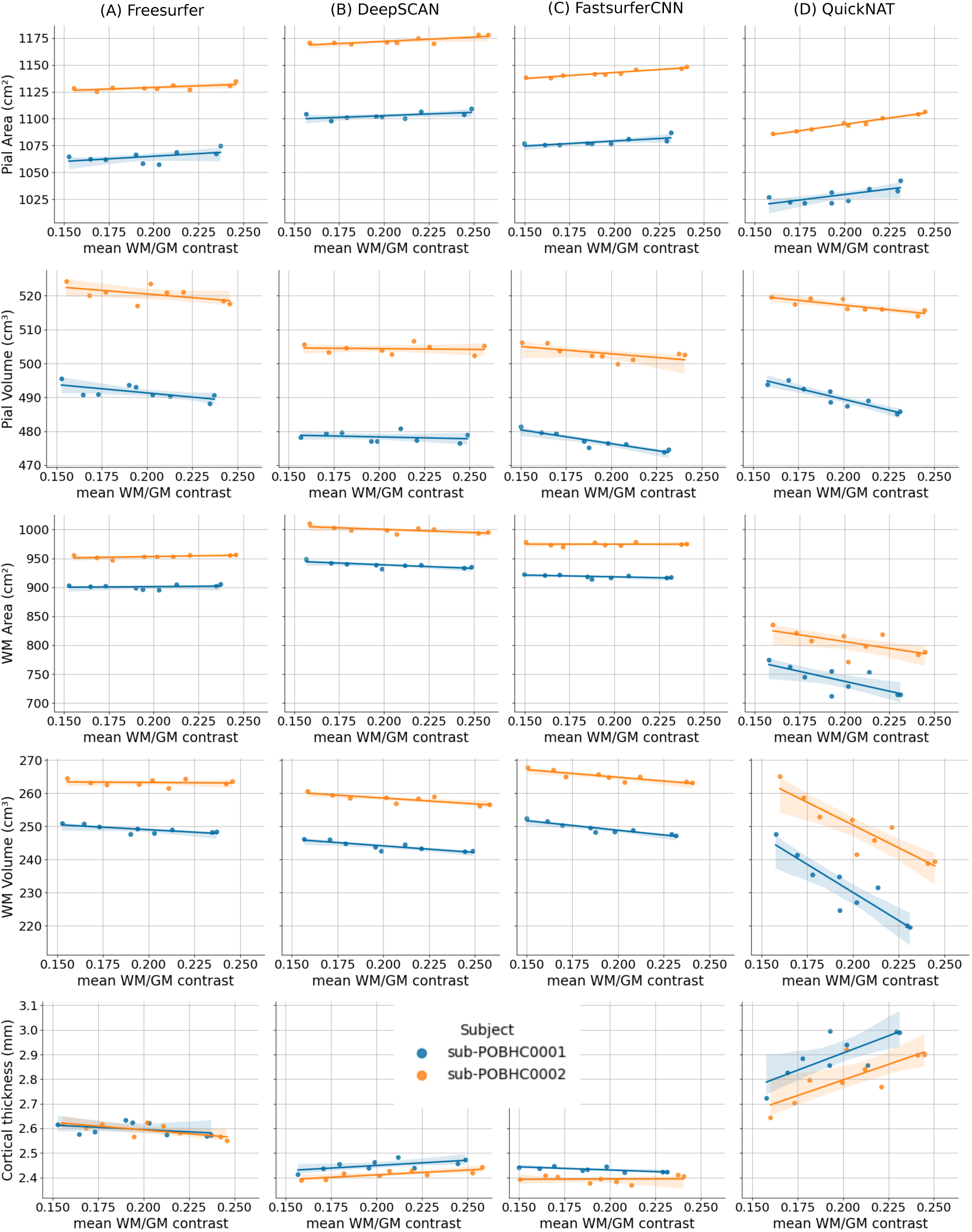
Correlation between the surface reconstruction and the morphological variables obtained from it with the MRI WM/GM contrast for (A) Freesurfer reconstruction and the fast surface pipeline based on (B) DeepSCAN, (C) FastsurferCNN and (D) QuickNAT. The average between left and right hemispheres were used to calculate the metrics for each data point.

**Table 4:**
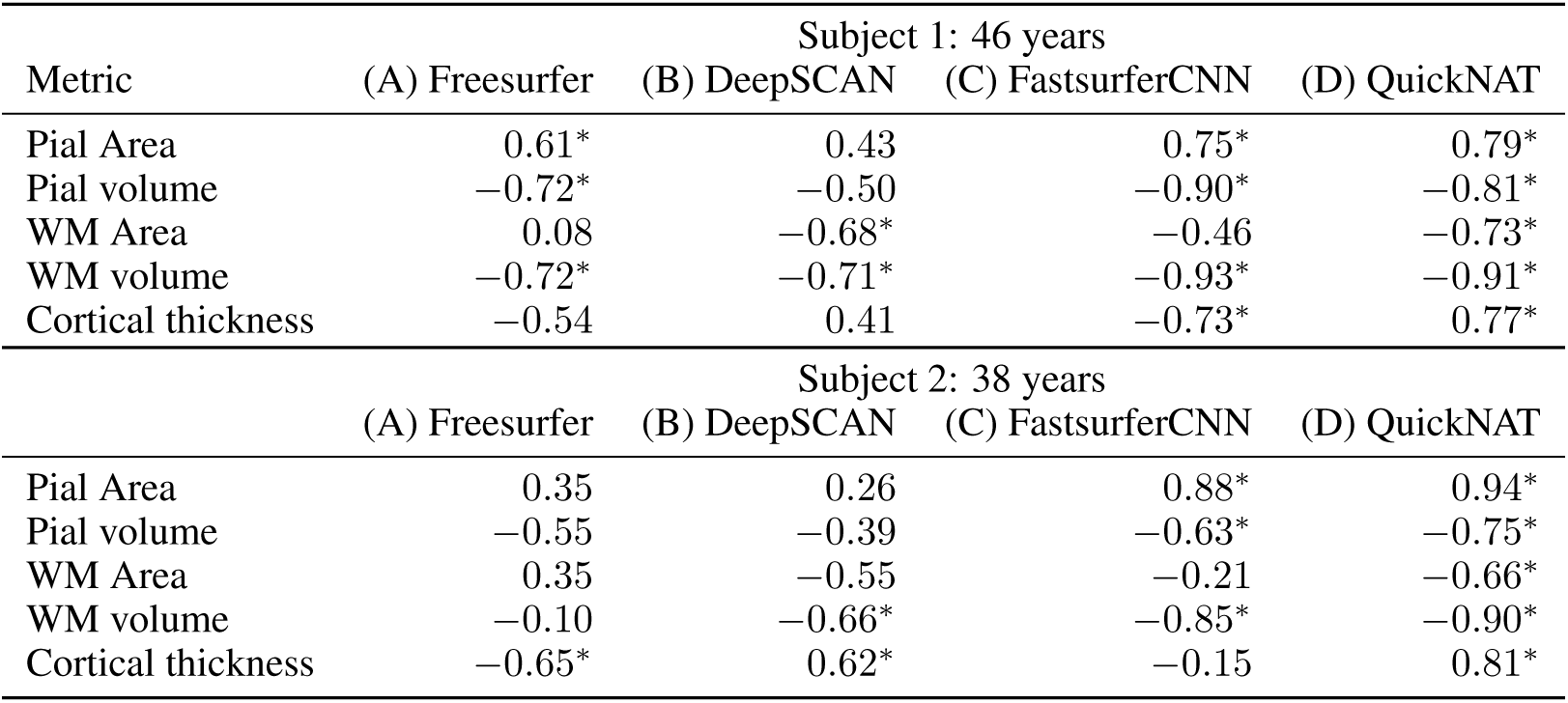
The Pearson correlation coefficient between the surface-based metrics and WM/GM contrast. The symbol * indicates the statistically significant results with *p <* 0.01.

| Metric | Subject 1: 46 years |  |  |  |
| --- | --- | --- | --- | --- |
|  | (A) Freesurfer | (B) DeepSCAN | (C) FastSurferCNN | (D) QuickNAT |
| Pial Area | 0.61* | 0.43 | 0.75* | 0.79* |
| Pial volume | -0.72* | -0.50 | -0.90* | -0.81* |
| WM Area | 0.08 | -0.68* | -0.46 | -0.73* |
| WM volume | -0.72* | -0.71* | -0.93* | -0.91* |
| Cortical thickness | -0.54 | 0.41 | -0.73* | 0.77* |

| Metric | Subject 2: 38 years |  |  |  |
| --- | --- | --- | --- | --- |
|  | (A) Freesurfer | (B) DeepSCAN | (C) FastSurferCNN | (D) QuickNAT |
| Pial Area | 0.35 | 0.26 | 0.88* | 0.94* |
| Pial volume | -0.55 | -0.39 | -0.63* | -0.75* |
| WM Area | 0.35 | -0.55 | -0.21 | -0.66* |
| WM volume | -0.10 | -0.66* | -0.85* | -0.90* |
| Cortical thickness | -0.65* | 0.62* | -0.15 | 0.81* |

### Benchmarking reconstruction pipelines

The synthetic dataset provides an expected value for the cortical thickness and ground-truth surfaces for the pial and white matter. Figure 7 shows the Hausdorff distance between the reconstructed surface and the ground-truth. The relative error obtained in the respective morphological variable is shown in Figure 8. The level of atrophy estimated by each reconstruction pipeline, including measurement with DiReCT instead of FreeSurfer, is presented in Figure 9. For all segmentation methods studied, the Freesurfer definition of cortical thickness fails to reproduce the synthetic atrophy. The DiReCT methodology improves the atrophy estimation showing a better performance when combined with DeepSCAN segmentation.

**Figure 7:**
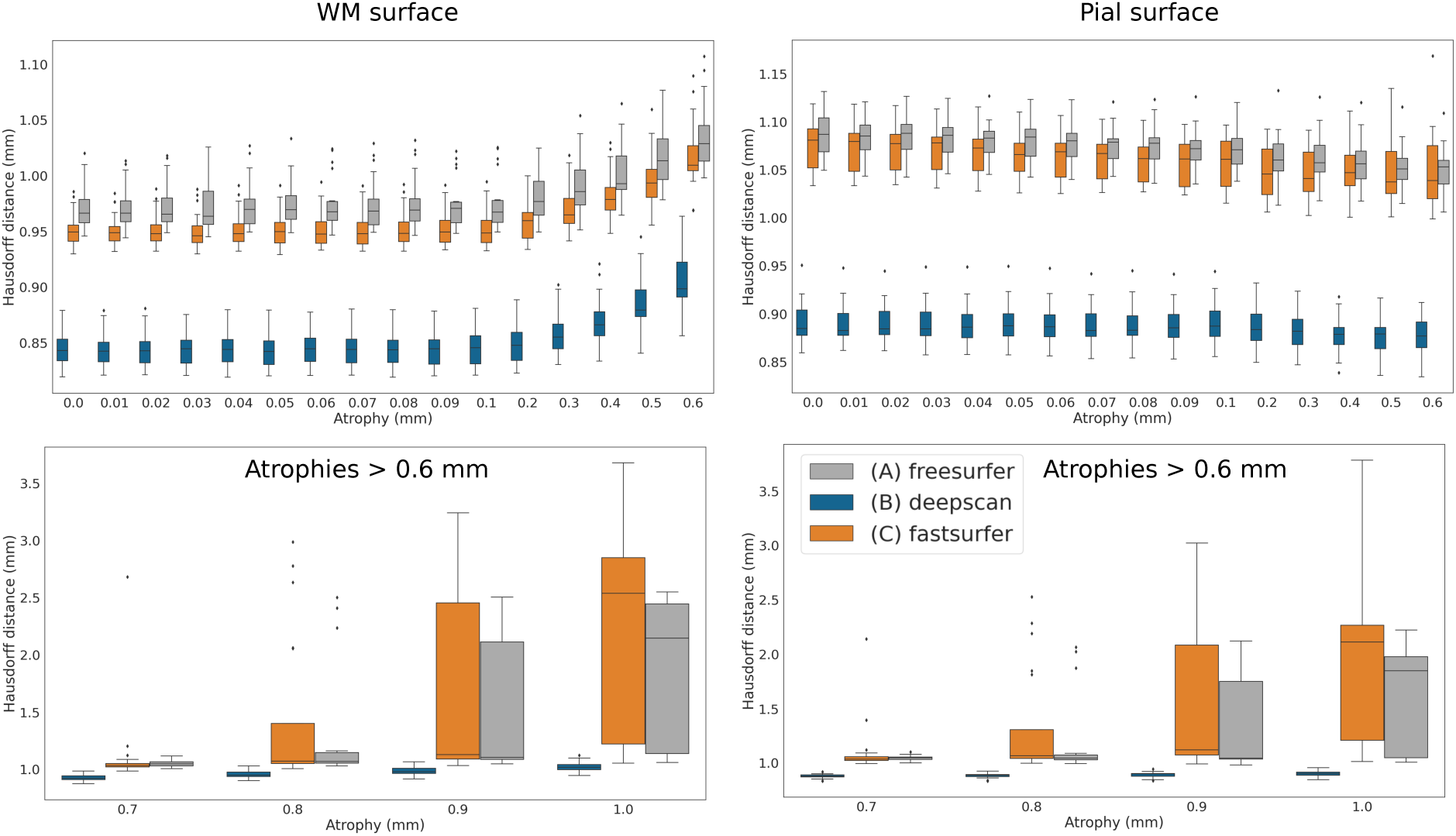
The Hausdorff distance calculated between the reconstructed surface and the ground-truth for the white matter surface (left) and the pial surface (right) using the (A) Freesurfer reconstruction and our fast surface pipeline based on (B) DeepSCAN and (C) FastsurferCNN to the synthetic ground truth surface.

**Figure 8:**
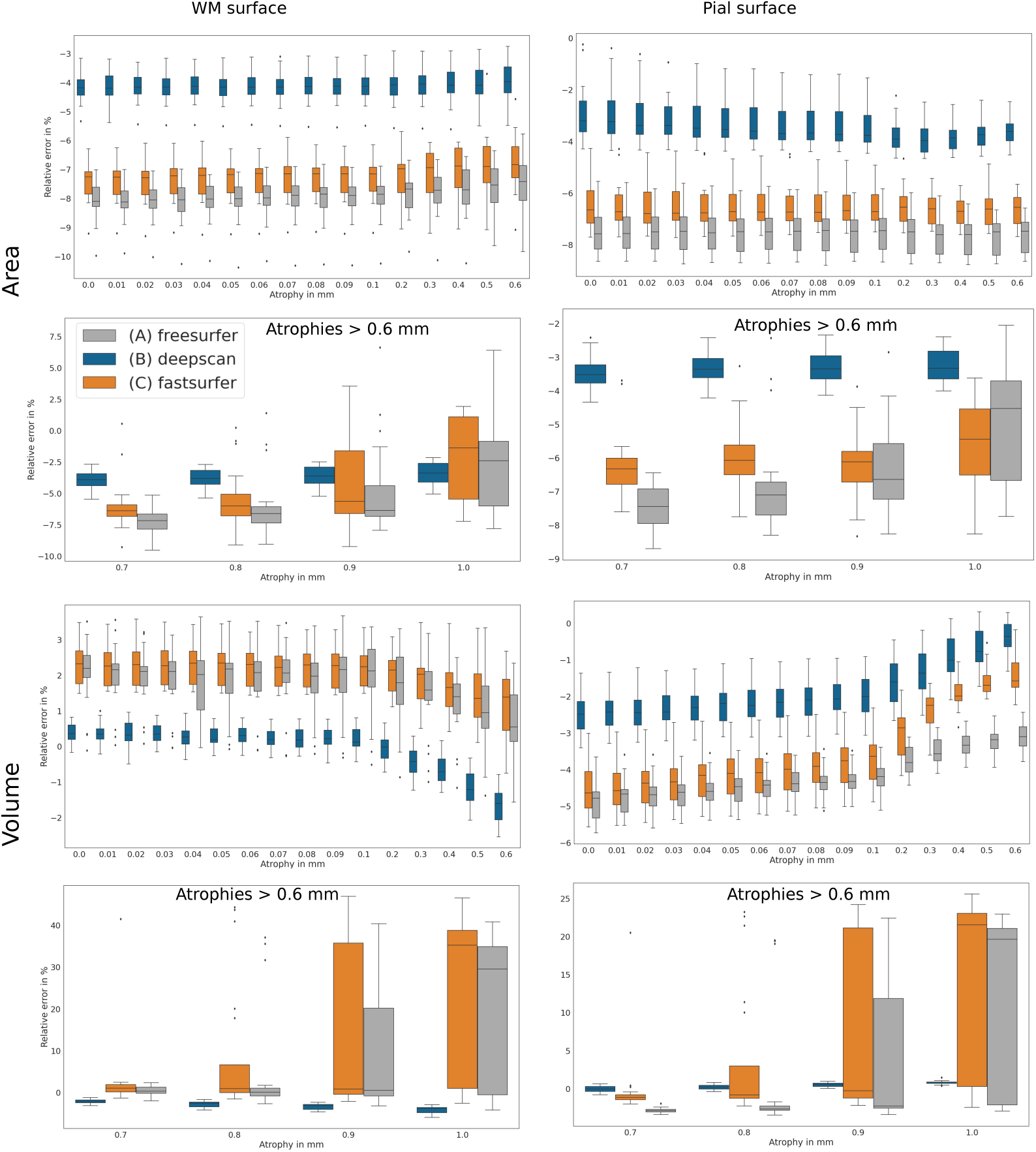
The relative error in the morphological variables derived from the surfaces reconstructed by (A) Freesurfer and the fast surface pipeline based on (B) DeepSCAN, (C) FastsurferCNN, using the synthetic ground-truth surface for comparison.

**Figure 9:**
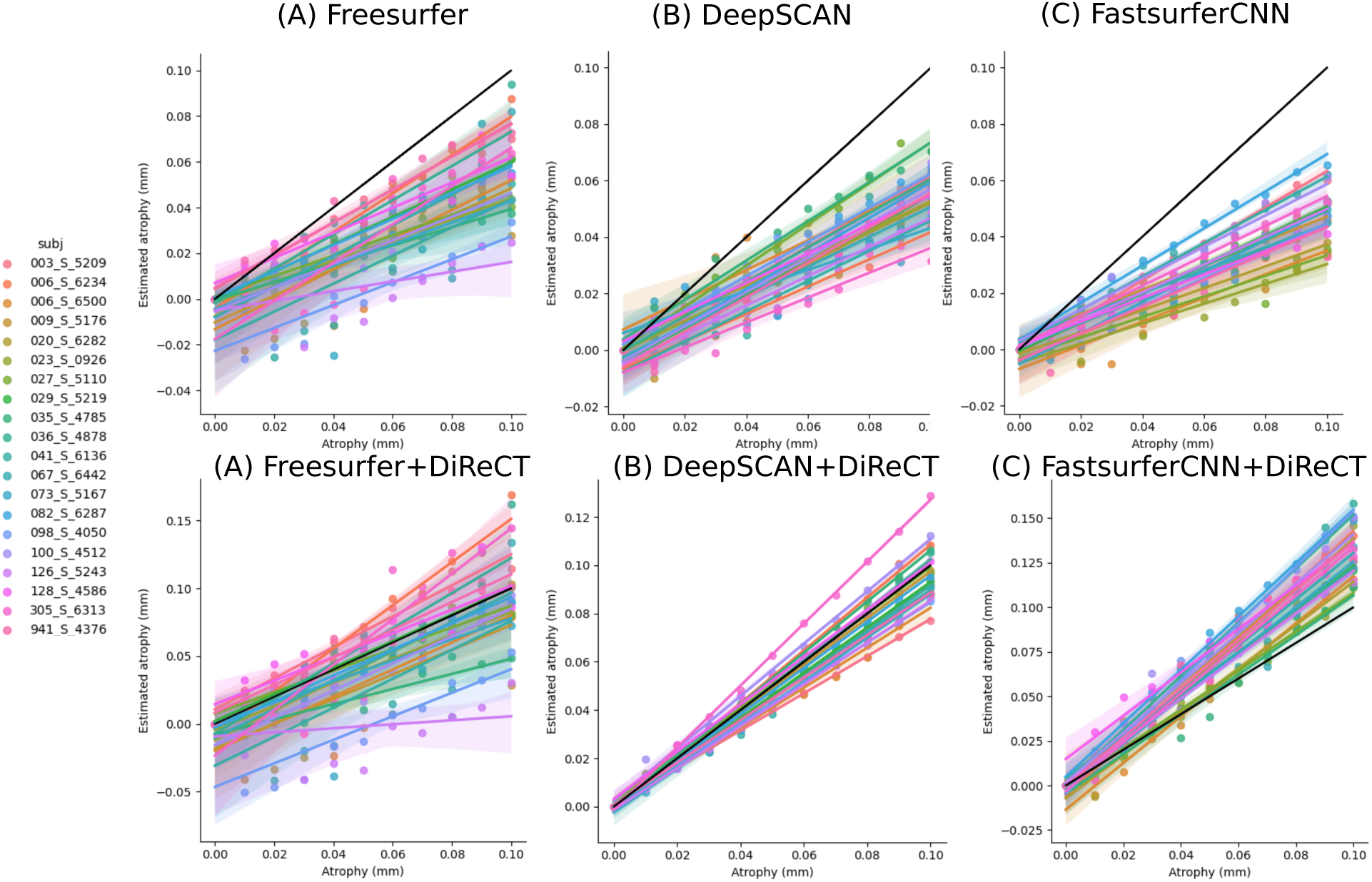
Estimated vs induced atrophy in the synthetic phantom, using both DiReCT and Freesurfer methodology [54]. The black line represents the identity.

## Discussion

There is evidence that DL-based methods are a promising approach for a fast whole-brain segmentation [25–28]. The results presented here extend previous work done in [28], not only adding information about the performance of another state of the art whole-brain segmentation model (DeepSCAN [27]) but also going beyond the voxel-based evaluation and dice coefficient comparison between the DL-based segmentation and Freesurfer’s silver standard. Using a methodology inspired by [28], we produced a fast surface reconstruction pipeline to be used with any parcellation of a structural MRI into cortical regions and WM. With a processing runtime of 11 minutes (Figure 2), this pipeline reflects the minimum amount of time required to generate a surface to be further manipulated according to the desired application.

The general overview presented in Figure 1 indicates that the surfaces obtained from the fast reconstruction pipeline are qualitatively equivalent to the ones obtained by Freesurfer. While there is no dissimilarity compared to Freesurfer in the (B) DeepSCAN based reconstructions, small deviations are apparent for (C) FastsurferCNN in the WM surface, only for the ABCD dataset. More significant differences are present for (D) QuickNAT in all reconstructions displayed. The inaccurate surface reconstructions are caused by a systematic underestimation of the WM volume by either missing regions and/or while delimiting the WM/GM boundary. Further examples are given in the supplementary material B and D.

Quantitatively, some differences between methodologies are revealed in both whole-hemisphere and region-wise analysis. For the whole-hemisphere, Figure 3 and the slope coefficients shown in Table 5 demonstrate that the surfaces based on (B) DeepSCAN are the closest to (A) Freesurfer pseudo ground-truth for all tested datasets. This observation is maintained in the region-wise analysis presented in Figure 4 for which (B) DeepSCAN shows less deviation than the other models. More, in the test-retest analysis shown in Figure 5, (B) DeepSCAN displays a level of reproducibility higher than both (A) Freesurfer and (C) FastsurferCNN, a result in agreement with the observation of [27]. The reconstructions based on (C) FastsurferCNN fail to reproduce Freesurfer in the ABCD dataset. As shown in the supplementary material A and D, these discrepancies are due to a systematic underestimation of the WM/GM boundary and are maintained even for the full Fastsurfer reconstruction. Although (C) FastsurferCNN metrics derived from the whole hemisphere are close to (A) Freesurfer pseudo ground-truth, region-wise there is significant differences. The reconstructions based on (D) QuickNAT have the most significant differences to the (A) Freesurfer pseudo ground-truth for all datasets due to an inaccurate WM/GM boundary delimitation.

Based on the level of robustness across a highly heterogeneous test dataset and the remarkably accelerated runtime of 11 minutes, considering the usage of the fast surface reconstruction pipeline based on combination (B) DeepSCAN+DiReCT becomes realistic for potential future clinical routine applications like diagnostic support [14–17]. In this context, the significant correlation between the resulting surface metrics with the WM/GM contrast of the used MRI sequence (see Figure 6 and Table 4, and already noticed in previous studies [57, 58]), is of great importance. The structural MRI acquired for clinical purposes presents an inherent variability regarding not only subjects and MRI machines but also sequence parameters. These effects should be taken into account in longitudinal and cross sectional comparisons. This stands as one of the major challenges to overcome for clinical routine applications.

The major difference in the morphometry derived from the fast surface reconstruction for both (B) DeepSCAN and (C) FastsurferCNN is observed for cortical thickness. As both pial and WM volumes reconstructed on the human dataset are in agreement with Freesurfer, meaning that they enclose similar regions, this suggests an instability on Freesurfer thickness estimation methodology, which is better delineated with the synthetic dataset. The synthetic dataset, as shown in Figures 7 and 8 revealed a general tendency of all reconstructions, including the silver standard Freesurfer, to underestimate the sizes of the pial and WM surface. Among the reconstructions, DeepSCAN showed a better performance in reconstructing the ground truth, presenting about 1.5-2 times less relative error than the others in terms of pial area, pial volume and WM area and nearly zero relative error in the WM volume estimation. The absence of outliers in the Hausdorff distance for the DeepSCAN pipeline, as illustrated in Figure 7, even when analyzing synthetic atrophies exceeding 0.7 mm, indicates that no implausible surfaces were detected for this pipeline. This contrasts with the results observed for Freesurfer and Fastsurfer. It is important to mention that, even though the synthetic dataset is a powerful tool to benchmark the reconstruction methods, it must be considered only as a *model* of the evolution of the cortical atrophy neurodegenerative. Values of synthetic atrophy as large as 0.5 mm or even more are rather unrealistic. However, the robustness in the reconstruction of these highly implausible brains can be interpreted as a indication of robustness of the pipeline itself. In terms of cortical thickness estimation, the Freesurfer methodology is unable to reproduce the introduced atrophy. For this particular metric, DiReCT provided results closer to the ground truth: this is in agreement with previously published results [27, 43].

In conclusion, our results suggests that, as long as the segmentation model is able provide a good WM delimitation, the fast surface reconstruction pipeline will reconstruct reliable surfaces with a 11 minute runtime. The methodology presented here is scalable to any segmentation map, opening the possibility of using updated versions of the current models, new DL-based segmentation, specialized models for contrast enhanced sequences or specific diseases [59–61]. In particular, contrast agnostic models such as [61] can be used to overcome the contrast dependency on the metrics. For the three state-of-the-art segmentation models examined in this work, the most robust pipeline across the human dataset and closer to the synthetic ground truth was based on (B) DeepSCAN segmentation and DiReCT estimation of cortical thickness. This reconstruction methodology stands out as a possibility to bridge the gap between research and clinical application with a reliable and robust morphometry tool.

## Data Availability

All data and codes used in the present study are public available.

## Acknowledgements

This work is supported by the Swiss National Science Foundation (SNF, grant 204593, “ScanOMetrics” project). C.R. and R.W. acknowledge other support by the SNF (grant CRSII5 180365, Swiss First project) and the NIH (grant R01NS107513, Enigma Parkinson). We are thankful to the research team and volunteers for participating in this research project. We also want to acknowledge the importance of the public data sharing initiative followed by the authors of ABDC, HCP, OASIS, Synthetic atrophy and Phantom of Bern studies. Without it, this work would not be possible.

## Supplementary Information

### A Full Fastsurfer reconstruction

The fast surface reconstructions based on (C) FastsurferCNN fail to reproduce Freesurfer in the ABCD dataset, see top row of Fig. 3. To ensure this effect is not due to our simplification of the pipeline, we have run the full Fastsurfer pipeline in a sub-sample of 100 random subjects. Figure 10 shows that the same dissimilarity is observed in the full Fastsurfer pipeline.

**Figure 10:**
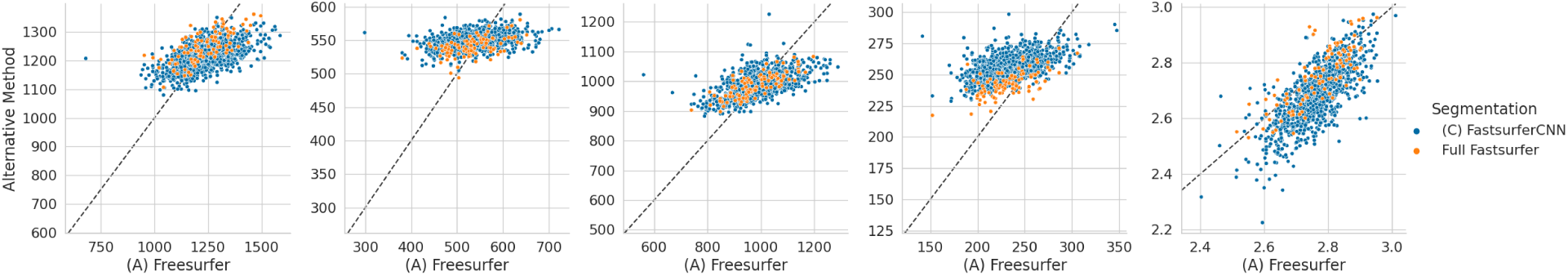
Comparison between the SBM of obtained using the full FastsurferCNN pipeline and the simplified pipeline with the standard (A) Freesurfer reconstruction.

Additionally, the surface obtained by the fast methodology does not displays significant regional differences to the Full Fastsurfer as shown in Figure 11. The median relative difference between the morphological variables derived from the full Fastsurfer and the fast reconstruction pipeline further indicates no effect on the final surface due to the pipeline simplification.

**Figure 11:**
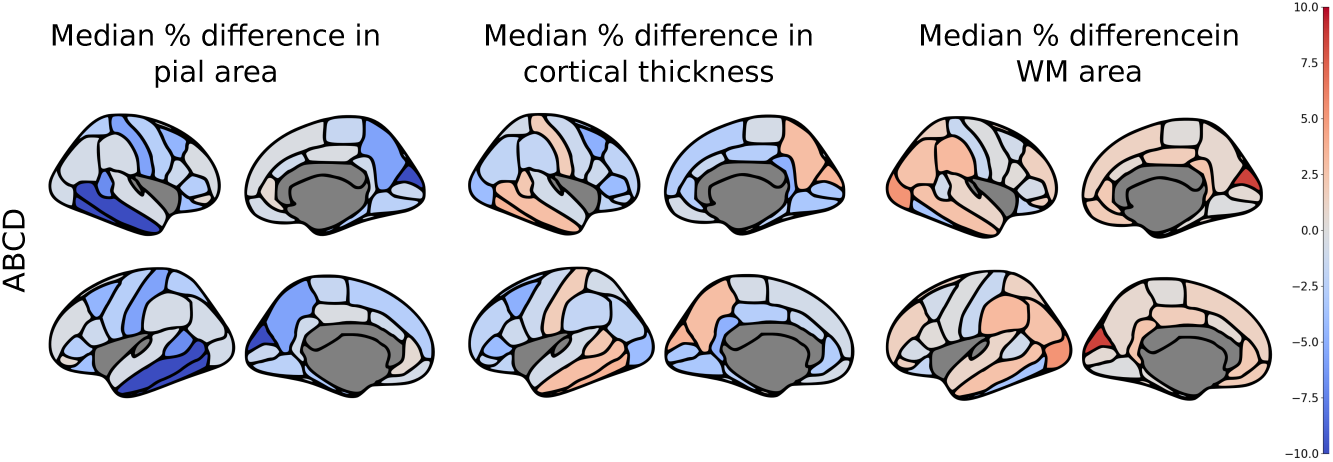
The Median relative difference between the surface based morphological variables derived from the full Fastsurfer reonstruction and the fast reconstruction pipeline.

### B General overview of the fast surface reconstruction

**Figure 12:**
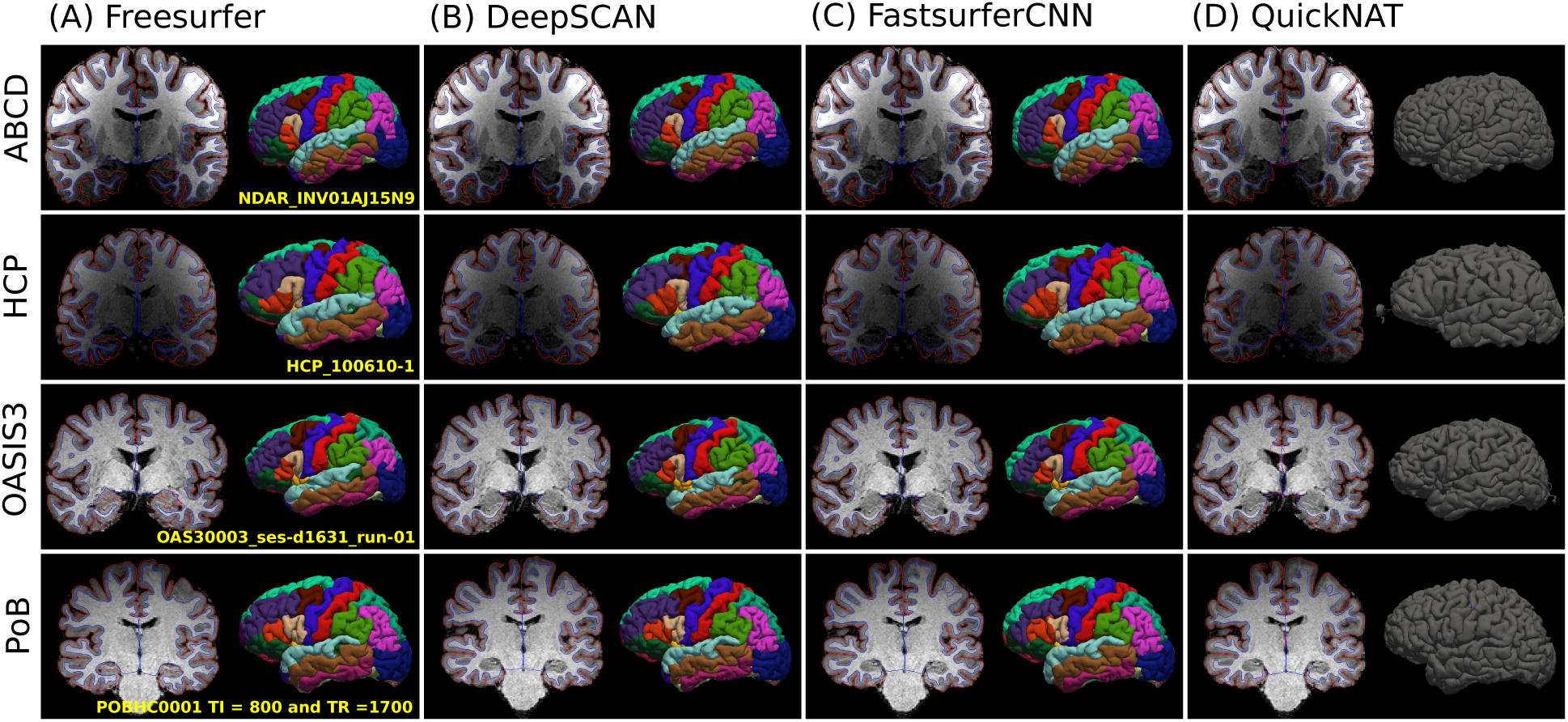
Example of the results obtained from the fast surface reconstruction pipeline based on (B) DeepSCAN, (C) FastsurferCNN and (D) QuickNAT segmentation in comparison with the standard (A) Freesurfer reconstruction. The raw MRI is displayed together with the reconstructed WM (blue) and pial surfaces (red) in coronal view for the same cross-section of the brain to facilitate a direct comparison between surfaces. Apart from occasional segmentation failures, the surfaces are similar except in the hippocampus, which is not enclosed inside the WM for (A) Freesurfer. The 3D pial surface showing the region labeling provided by each DL-based segmentation model allows a neuro-anatomical comparison with the DK Atlas. The subjects are identified for reproducibility and the anatomical visualization was kept consistent to facilitate comparison between reconstructions. We retained the same coloring and naming presented in [51].

### C Table with the linear regression coefficients

**Table 5:** Slope and offset obtained by the linear regression between the Freesurfer’s metrics and the one obtained by the alternative methods (see Figure 3).

| Dataset | Metric | (B) DeepSCAN | (C) FastSurferCNN | (D) QuickNAT |
| --- | --- | --- | --- | --- |
| ABCD | Pial area | $(0.92 \pm 0.01)$<br>$(5 \pm 1) \text{ cm}^2$ | $(0.24 \pm 0.01)$<br>$(91 \pm 1) \text{ cm}^2$ | $(0.88 \pm 0.02)$<br>$(8 \pm 3) \text{ cm}^2$ |
| | Pial volume | $(0.92 \pm 0.01)$<br>$(17 \pm 6) \text{ cm}^3$ | $(0.07 \pm 0.01)$<br>$(510 \pm 5) \text{ cm}^3$ | $(0.96 \pm 0.04)$<br>$(4 \pm 200) \text{ cm}^3$ |
| | WM area | $(0.88 \pm 0.01)$<br>$(7 \pm 1) \text{ cm}^2$ | $(0.25 \pm 0.01)$<br>$(74 \pm 1) \text{ cm}^2$ | $(0.80 \pm 0.03)$<br>$(3 \pm 4) \text{ cm}^2$ |
| | WM volume | $(0.96 \pm 0.01)$<br>$(11 \pm 3) \text{ cm}^3$ | $(0.19 \pm 0.01)$<br>$(211 \pm 3) \text{ cm}^3$ | $(0.92 \pm 0.03)$<br>$(8 \pm 85) \text{ cm}^3$ |
| | Cortical thickness | $(0.80 \pm 0.02)$<br>$(0.35 \pm 0.06) \text{ mm}$ | $(0.93 \pm 0.03)$<br>$(0.1 \pm 0.1) \text{ mm}$ | $(0.70 \pm 0.10)$<br>$(0.9 \pm 0.4) \text{ mm}$ |
| HCP | Pial area | $(0.98 \pm 0.01)$<br>$-(1.3 \pm 0.3) \text{ cm}^2$ | $(1.00 \pm 0.01)$<br>$-(0.9 \pm 0.3) \text{ cm}^2$ | $(0.95 \pm 0.01)$<br>$(5 \pm 1) \text{ cm}^2$ |
| | Pial volume | $(0.96 \pm 0.01)$<br>$-(6 \pm 1) \text{ cm}^3$ | $(0.97 \pm 0.01)$<br>$(5 \pm 2) \text{ cm}^3$ | $(0.92 \pm 0.02)$<br>$(50 \pm 6) \text{ cm}^3$ |
| | WM area | $(0.99 \pm 0.01)$<br>$-(2.1 \pm 0.3) \text{ cm}^2$ | $(1.00 \pm 0.01)$<br>$-(0.1 \pm 0.4) \text{ cm}^2$ | $(0.80 \pm 0.01)$<br>$(7 \pm 1) \text{ cm}^2$ |
| | WM volume | $(1.00 \pm 0.01)$<br>$(2.8 \pm 0.7) \text{ cm}^3$ | $(0.97 \pm 0.01)$<br>$(10 \pm 1) \text{ cm}^3$ | $(0.80 \pm 0.01)$<br>$(43 \pm 3) \text{ cm}^3$ |
| | Cortical thickness | $(0.79 \pm 0.02)$<br>$(0.33 \pm 0.04) \text{ mm}$ | $(0.78 \pm 0.03)$<br>$(0.43 \pm 0.08) \text{ mm}$ | $(0.78 \pm 0.07)$<br>$(0.81 \pm 0.2) \text{ mm}$ |
| OASIS3 | Pial area | $(0.98 \pm 0.01)$<br>$(0.3 \pm 0.4) \text{ cm}^2$ | $(0.98 \pm 0.01)$<br>$(0.8 \pm 0.4) \text{ cm}^2$ | $(0.99 \pm 0.02)$<br>$(1 \pm 10) \text{ mm}^2$ |
| | Pial volume | $(0.94 \pm 0.01)$<br>$(6 \pm 2) \text{ cm}^3$ | $(0.97 \pm 0.01)$<br>$(12 \pm 2) \text{ cm}^3$ | $(1.02 \pm 0.02)$<br>$-(5 \pm 5) \text{ cm}^3$ |
| | WM area | $(0.94 \pm 0.01)$<br>$(2.7 \pm 0.4) \text{ cm}^2$ | $(0.94 \pm 0.01)$<br>$(3.6 \pm 0.5) \text{ cm}^2$ | $(0.83 \pm 0.03)$<br>$(0 \pm 1) \text{ cm}^2$ |
| | WM volume | $(0.98 \pm 0.01)$<br>$(7 \pm 1) \text{ cm}^3$ | $(0.99 \pm 0.01)$<br>$(9 \pm 1) \text{ cm}^3$ | $(0.91 \pm 0.02)$<br>$(1 \pm 3) \text{ cm}^3$ |
| | Cortical thickness | $(0.70 \pm 0.01)$<br>$(0.51 \pm 0.04) \text{ mm}$ | $(0.78 \pm 0.01)$<br>$(0.44 \pm 0.04) \text{ mm}$ | $(0.82 \pm 0.06)$<br>$(0.8 \pm 0.1) \text{ mm}$ |

### D General overview of the WM segmentation

As a supplement to Figure 3, a general overview of the WM segmentation provided by the three DL-based methods is shown in Figures 13 to 15. The representative subjects were chosen using the first (close to Freesurfer), second (median behavior) and third (far from Freesurfer) quartiles of the percentage difference between the WM volume reconstructed and Freesurfer’s silver standard.

**Figure 13:**
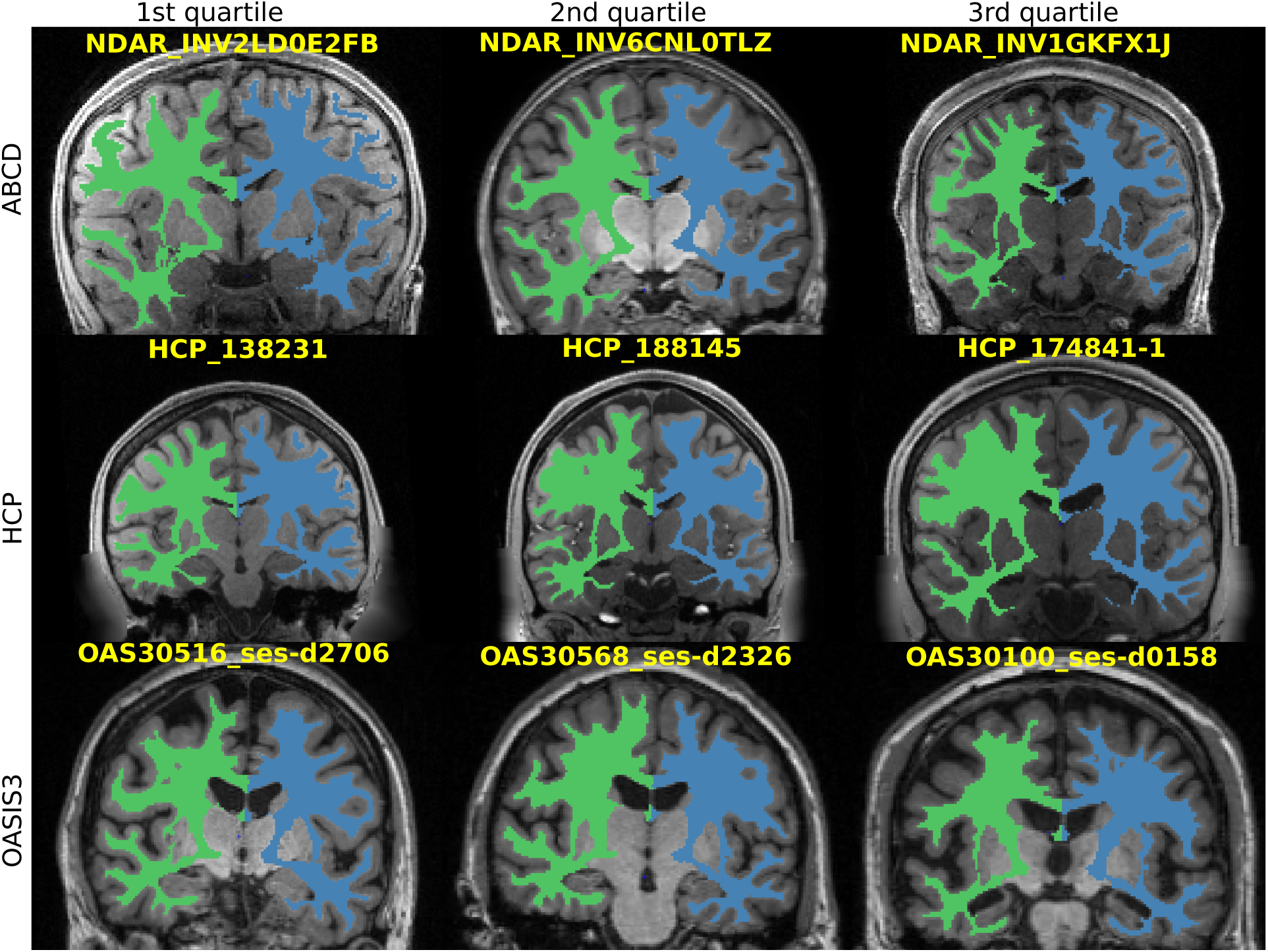
Examples of the WM segmentation provided by (D) QuickNAT.

There is a systematic underestimation of the WM either missing regions and/or in the WM/GM boundary delimitation. This explains the most significant differences observed when comparing (D) QuickNAT with Freesurfer reconstruction, for all datasets. This systematic tendency to underestimate the WM volume can also be seen on (C) FastsurferCNN prediction for the ABCD dataset only. Figures 14 and 15 shows a general overview of the WM segmentation for both (B) DeepSCAN and (C) FastsurferCNN.

**Figure 14:**
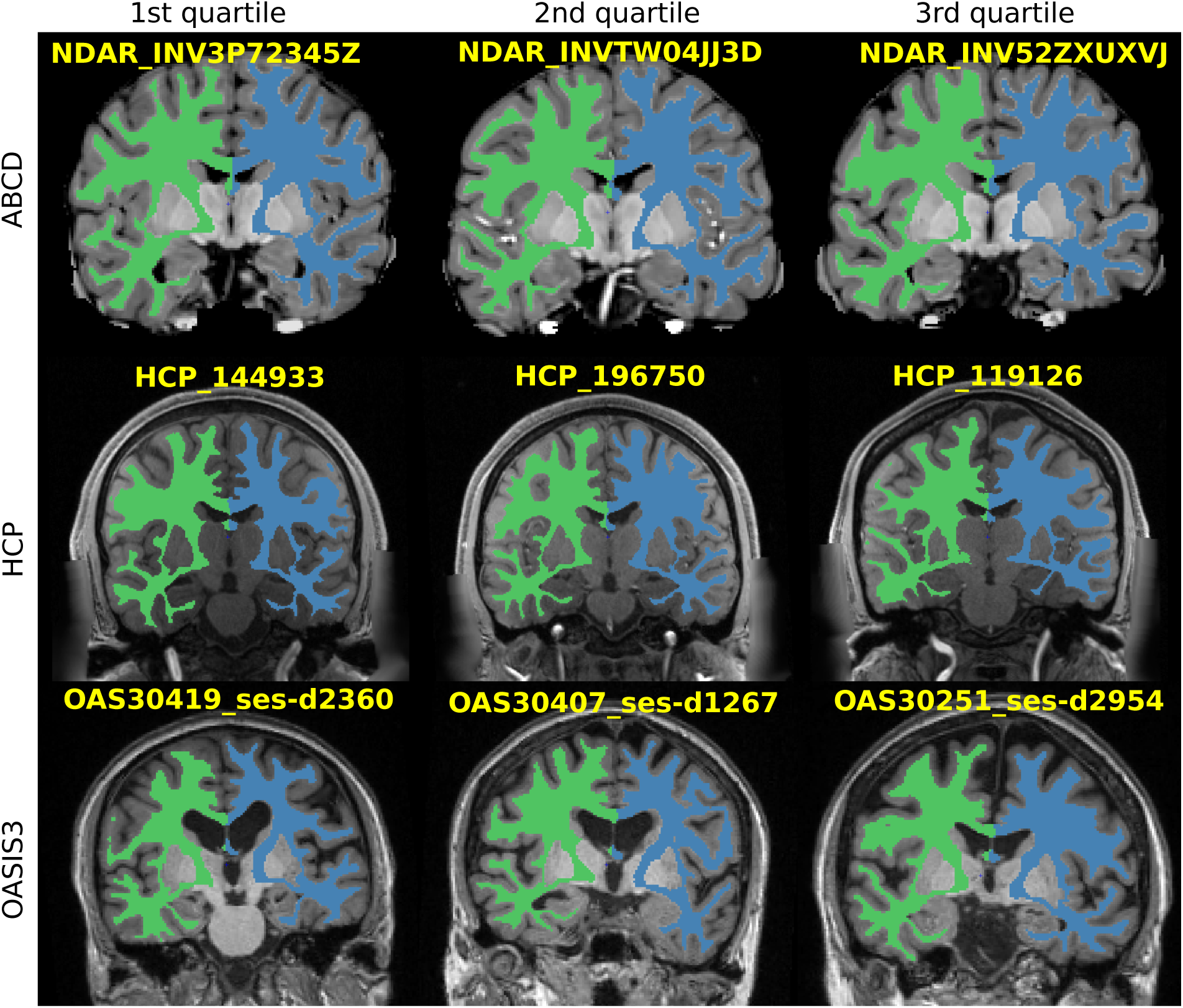
Examples of the WM segmentation provided by (C) FastsurferCNN.

**Figure 15:**
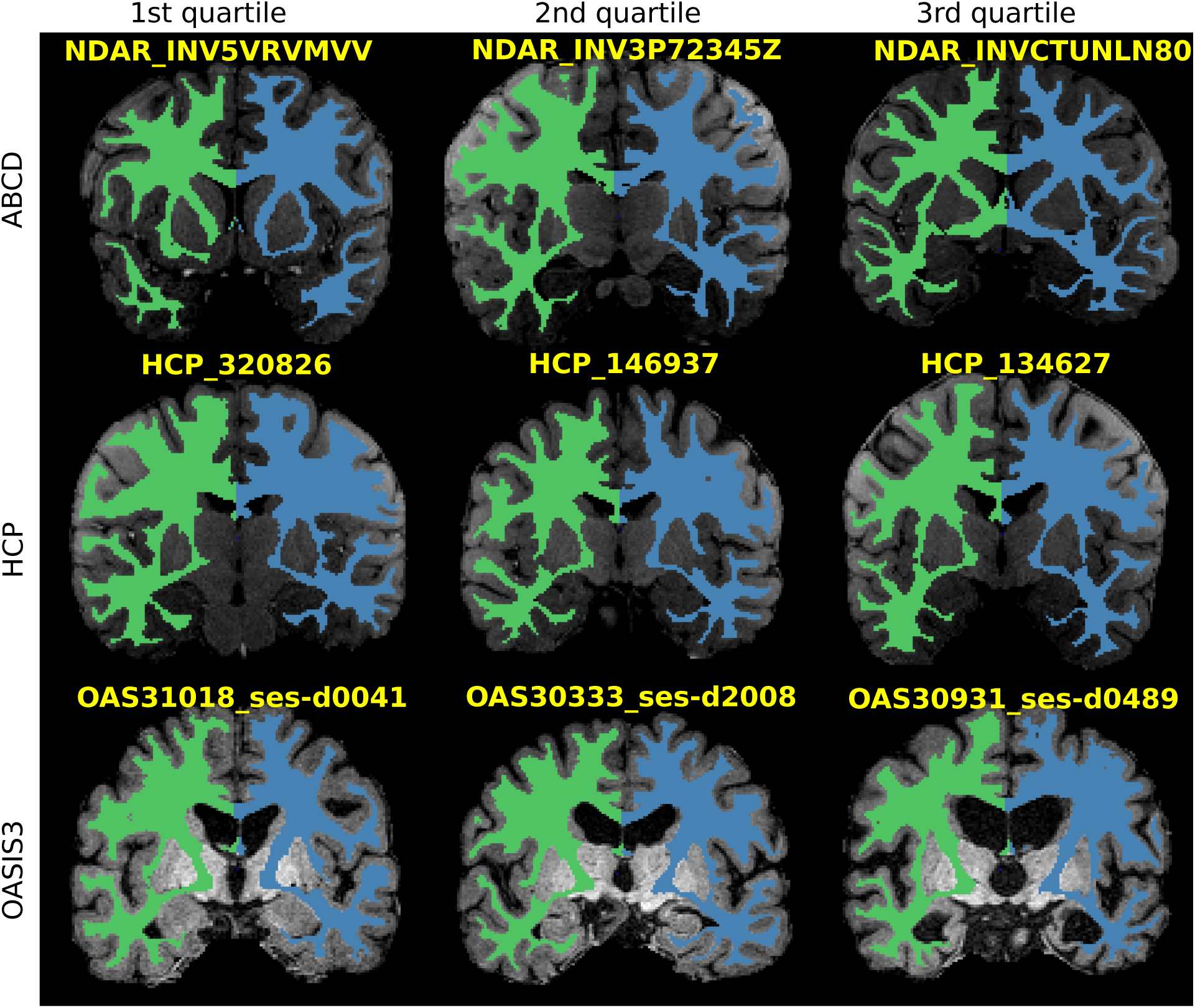
Examples of the WM segmentation provided by both (B) DeepSCAN.

